# Comparing Sexual Network Mean Active Degree Measurement Metrics among Men who have Sex with Men

**DOI:** 10.1101/2022.02.11.22270855

**Authors:** Christina Chandra, Martina Morris, Connor Van Meter, Steven M. Goodreau, Travis Sanchez, Patrick Janulis, Michelle Birkett, Samuel M. Jenness

## Abstract

**Background:** Mean active degree is an important proxy measure of cross-sectional network connectivity commonly used in HIV/STI epidemiology research. No current studies have compared measurement methods of mean degree using cross-sectional surveys for men who have sex with men (MSM) in the United States.

**Methods:** We compared mean degree estimates based on reported ongoing main and casual sexual partnerships (*current method*) against dates of first and last sex (*retrospective method*) from 0–12 months prior to survey date in ARTnet, a cross-sectional survey of MSM in the U.S. (2017–2019). ARTnet collected data on the number of sexual partners in the past year but limited reporting on details used for calculating mean degree to the 5 most recent partners. We used linear regression to understand the impact of truncated partnership data on mean degree estimation.

**Results:** Retrospective method mean degree systematically decreased as the month at which it was calculated increased from 0–12 months prior to survey date. Among participants with >5 partners in the past year compared to those with ≤5, the average change in main degree between 12 and 0 months prior to survey date was −0.05 (95% CI: −0.08, −0.03) after adjusting for race/ethnicity, age, and education. The adjusted average change in casual degree was −0.40 (95% CI: −0.45, −0.35).

**Conclusions:** The retrospective method underestimates mean degree for MSM in surveys with truncated partnership data, especially for casual partnerships. The current method is less prone to bias from partner truncation when the target population experiences higher cumulative partners per year.

**Summary:** Survey designs can lead to potential bias, such as underestimation, in the measurement of mean active degree in sexual networks of men who have sex with men.

## INTRODUCTION

In the United States, men who have sex with men (MSM) experience a disproportionate burden of HIV and bacterial sexually transmitted infections (STIs). In 2018, MSM accounted for 66% of all new HIV diagnoses, 54% of primary and secondary syphilis cases, and 43% of gonorrhea cases.^1,2^ Young, non-white MSM are particularly affected by HIV/STIs despite evidence that individual risk behaviors are not different between Black and white MSM.^3^ Differences in sexual network connectivity, the mechanistic pathway for HIV/STI transmission, may explain these disparities among MSM in the U.S.^4,5^ Networks are also essential for deploying prevention tools, such as HIV PrEP or STI partner services.^6,7^

The potential effectiveness of a network-informed HIV/STI public health response depends on good empirical metrics: accurate and unbiased estimates of observable behaviors that determine the unobservable network connectivity. One common metric is active degree: the count of current, ongoing partners at a point in time, which may be summarized as the mean active degree across nodes (persons) at the population level. It is mathematically established and intuitive that network connectivity rises as mean degree increases, though the relation is non-linear.^7^ Active degree also forms the definitional basis of partnership concurrency (active degree of two or more). Modeling has demonstrated that higher mean active degree and higher prevalence of concurrency create network conditions that lead to more rapid and pervasive HIV/STI spread within networks.^5,8^ Even small changes in mean active degree can have a substantial impact on network connectivity, and subsequently epidemic persistence or elimination, due to the non-linear threshold effects.^9^ For this reason, accurate measurement of mean active degree is needed to assess epidemic potential.

UNAIDS focused on this problem in a reference group meeting in 2009, leading to published recommendations for measuring active degree and calculating point prevalence of concurrency as an indicator for monitoring national HIV epidemics.^10^ Approaches they considered were: 1) active degree as measured on the day of survey (“current method”); 2) using reported dates of first and last sex with each partner during the last year (“retrospective method”). UNAIDS recommended the latter. **Figure 1** provides a schematic of how the current and retrospective methods are used to calculate mean degree with hypothetical data.

**Figure 1.**
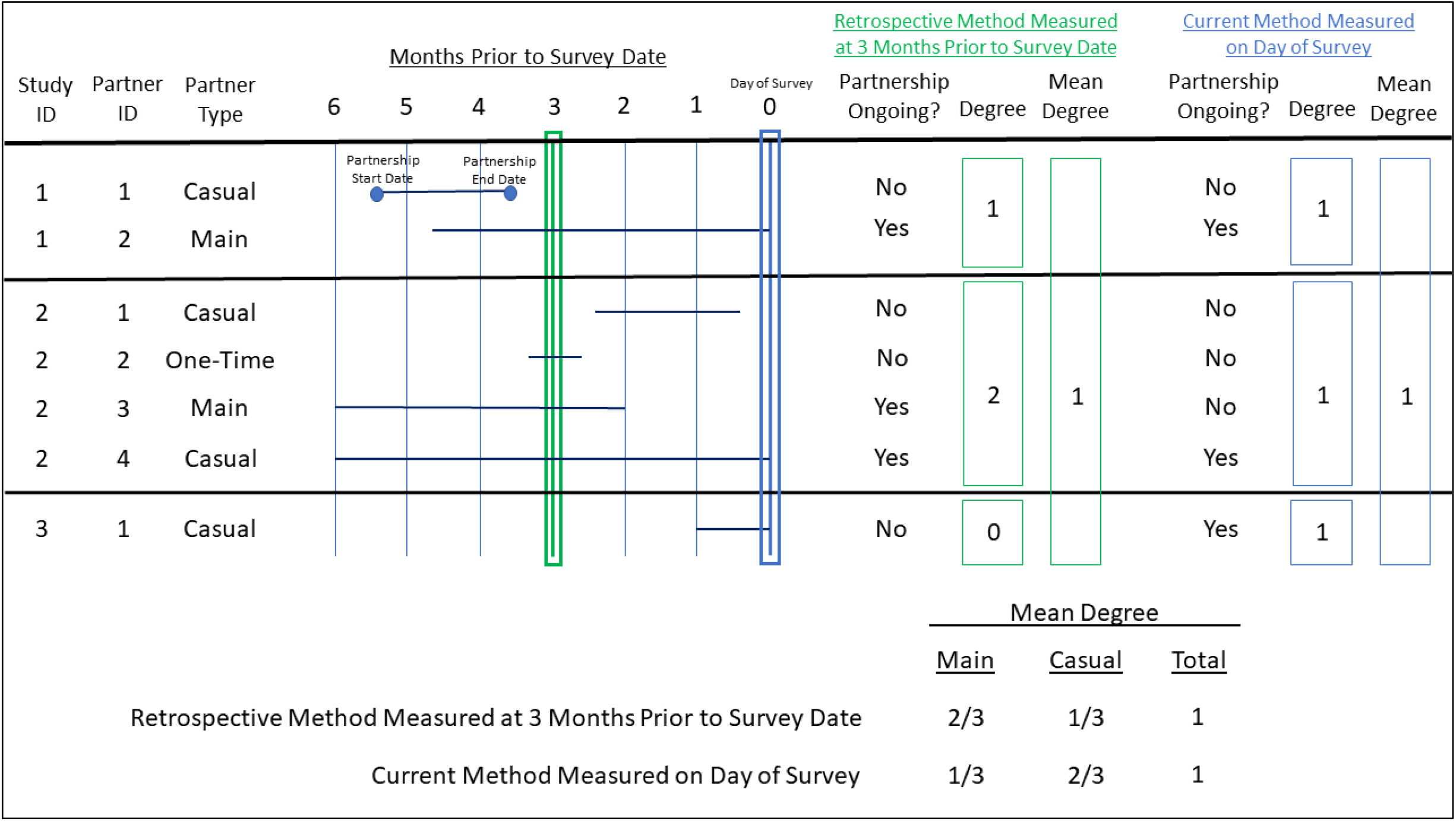
Schematic representation of how mean degree is calculated using hypothetical sexual partnership data with the retrospective method when measuring mean degree at three months prior to survey date and the current method where mean degree is measured on survey date. Mean degree can be calculated for main and casual sexual partnerships, and one-time partnerships are not included in the calculation by definition as they are not ongoing sexual partnerships.

The goal of the current method is to measure the person’s ongoing sexual partnerships on the day of the survey. This can be done on a partner-by-partner basis after enumerating relevant partners (e.g., “Do you expect to have sex with this person again?”), or with a single summary question (e.g., “With how many people do you currently have an active sexual relationship?”). Active degree is measured as the sum of ongoing partnerships reported. This approach does not depend on recall of events or dates and can be asked in a single question if time and respondent burden are concerns. The drawback is that respondents must accurately predict whether a partnership will continue. While that may be reasonable for populations characterized by having few long-term stable partnerships, it may fail in populations with frequent short-term casual partnerships.^11^

With the retrospective method, partners are enumerated for some period (e.g., all in the last year) and the partnership’s date of first and last sex are recorded. The partnership intervals are then evaluated for active overlap at specified time points prior to the survey date. The UNAIDS group recommended using 6 months prior to survey date to get a point estimate of the active degree. This method does not require the respondent to predict whether current partnerships will continue, but it requires accurate recollection of the dates of first/last sex. For surveys that ask for partnership dates for a limited number of partners (e.g., 5 most recent partners), participants with more sex partners may reach this limit before the specified number of months prior to survey date. For example, if a participant’s 5 most recent partnerships occurred within 3 months of the survey date, any additional partnerships at 6 months prior to the survey date would not be counted. The higher the cumulative number of partners over time, the more potential there is for downward bias in active degree as you go back in time.

While both methods have been used in surveys of MSM in the U.S.,^12–15^ there have been no comparative studies of these methods for MSM. Compared to heterosexual men and women, active degree is often higher among MSM, and more heterogenous, reflecting different partnership types of varying durations and typologies.^16^ Furthermore, the biases from truncated partnership data in cross-sectional studies (also known as fixed choice design and right-censoring of degree) has been well-documented in the field of network science.^17–19^ It would be useful to understand how these measurement approaches perform for MSM with this survey design.

In this study, we compared estimates using the current and the retrospective methods among U.S. MSM in a survey that allows for estimation with both methods and collects truncated partnership data. We aimed to understand if partnership data truncation was the main source of bias for the differences between the current and retrospective method, even after controlling for demographic variables that may be correlated with both truncation of partnership data and other mechanisms for bias in mean degree estimates.

## METHODS

### Study Design

We used data from ARTnet, a cross-sectional web-based study of MSM in the U.S. conducted between 2017 and 2019.^20^ ARTnet recruited participants through the American Men’s Internet Survey (AMIS), an ongoing study about MSM sexual health.^21^ ARTnet eligibility criteria included age between 15 and 65 years, cisgender male identity, male sex at birth, and having ever had sex with a male partner. ARTnet recruited AMIS participants and collected data in two waves: July 2017–February 2018 and September 2018–January 2019. Participants were deduplicated within and across waves. For participants with more than one survey record, we retained the most recent survey record from either wave. The Emory University Institutional Review Board approved this study.

### Measures

ARTnet collected information on egocentric sexual network data, recent sexual behavior, and utilization of HIV and STI prevention services. Participants were asked to provide information on up to 5 of their most recent male sexual partners within the last 12 months. Participants reported on type of partnership (main, casual, or one-time), dates of first and last sex by month and year, and sexual activity (e.g., condomless anal sex). Participants also answered the question, “Is this relationship with this partner active and ongoing?” If participants answered “Yes” but reported sex only once with the partner, then the partnership was considered a one-time partnership and not factored into mean degree calculations. We ran a sensitivity analysis to understand how an alternative definition where these partnerships were considered ongoing, casual partnerships and how imputing “Don’t know” answers to “Yes” impacted the mean degree of casual partnerships.

Our main exposure was the truncation of partnership data from limiting partnership information to the 5 most recent partners. ARTnet also asked participants for the total number of male sexual partners within the last 12 months, although details such as dates of first/last sex were not collected on all partners. The main exposure of partnership truncation was operationalized by a binary variable that split participants by whether they had 5 or fewer partners, including main, casual, or one-time, or more than 5 partners in the past 12 months. Those with more than 5 male partners in the past year are considered to have provided truncated partnership data. Our outcome of interest was a summary measure of active degree (henceforth, degree): the mean degree across all participants at a specified time point. This measure did not include one-time partnerships, as they are not ongoing. We calculated mean degree using the current method and compared it to mean degree calculated using the retrospective method at specified months prior to the survey date, ranging from 0 (current study month) to 12 months. We refer to the months prior to the survey date as monthly “offsets” (e.g., 6 months prior to the survey date is a 6-month offset). Since participants reported only month and year for dates of first/last sex in ARTnet, we randomly imputed days within these months. We conducted a sensitivity analysis to understand how date imputation impacted mean degree estimates from the retrospective method.

### Statistical Analysis

For descriptive analyses, we plotted mean degree by both current and retrospective methods by the primary exposure variable of having 5 or fewer or more than 5 partners. Our main outcome was the difference in degree between 12- and 0-month offsets. This difference provides a measure of stability of the retrospective method across offset months. Bivariable linear regression was used to estimate that 12-month difference and variations in that difference by the primary exposure variable and selected demographic variables (e.g., race/ethnicity, age, geography, education, income) that may also be correlated with bias in mean degree estimates. In multiple linear regression, we investigated whether variations in the 12- and 0-month degree difference were caused by the truncation of partnership data, after controlling for race, age, and education, which we hypothesized as associated with the primary exposure and the outcome. All analyses were conducted in R 4.2.0.^22^ Analysis scripts are provided in a GitHub repository (https://github.com/EpiModel/Mean-Degree-Analysis).

## RESULTS

Of the 4,904 MSM who completed the ARTnet study, most were non-Hispanic white, less than 35 years old, completed college or above, and had an annual household income of at least $40,000 (**Table 1**). A total of 16,198 partnerships were reported by the participants. Of these, 7,602 (46.9%) were one-time partnerships, 5,978 (36.9%) were casual partnerships, and 2,618 (16.2%) were main partnerships. On the day of survey, 5,875 (68.3%) of main and casual partnerships were reported as ongoing. Overall, 1,962 (40.0%) of participants had more than 5 partners (main, casual, and one-time) in the past 12 months.

**Table 1.** Individual- and Sexual Partnership-level Characteristics of Participants in ARTnet Study (2017–2019) of Men who have Sex with Men in the U.S.

| **Individual** | **N** | **%** |
| --- | --- | --- |
| **Total Sample** | 4904 | 100.0 |
| **Age (Mean and SD)** | 36.5 | 14.2 |
| **Age Category** |  |  |
| 15–24 | 1324 | 27.0 |
| 25–34 | 1268 | 25.9 |
| 35–44 | 694 | 14.2 |
| 45–54 | 833 | 17.0 |
| 55–65 | 785 | 16.0 |
| **Race/Ethnicity** |  |  |
| Black (Non-Hispanic) | 266 | 5.4 |
| Hispanic | 676 | 13.8 |
| Other (Non-Hispanic) | 439 | 9.0 |
| White (Non-Hispanic) | 3523 | 71.8 |
| **Residence in Census Region** |  |  |
| Northeast | 882 | 18.0 |
| Midwest | 994 | 20.3 |
| South | 1782 | 36.3 |
| West | 1246 | 25.4 |
| **Residence in Census Division** |  |  |
| New England | 250 | 5.1 |
| Middle Atlantic | 632 | 12.9 |
| East North Central | 698 | 14.2 |
| West North Central | 296 | 6.0 |
| South Atlantic | 1062 | 21.7 |
| East South Central | 222 | 4.5 |
| West South Central | 498 | 10.2 |
| Mountain | 404 | 8.2 |
| Pacific | 842 | 17.2 |
| **Education** |  |  |
| High school or below | 616 | 12.6 |
| Some college | 1519 | 31.1 |
| College and above | 2742 | 56.2 |
| **Annual Household Income** |  |  |
| \$0 to \$19,999 | 605 | 13.7 |
| \$20,000 to \$39,999 | 867 | 19.6 |
| \$40,000 to \$74,999 | 1258 | 28.5 |
| \$75,000 or more | 1686 | 38.2 |
| **Number of Male Partners in the Past 12 Months** |  |  |
| ≤ 5 Partners | 2939 | 59.9 |
| > 5 | 1962 | 40.0 |
| **Sexual Partnerships<sup>a</sup>** | **N** | **%** |
| **Total Sample** | 16198 | 100.0 |
| **Sexual Partnership Type** |  |  |
| Main | 2618 | 16.2 |
| Casual | 5978 | 36.9 |
| One-time | 7602 | 46.9 |
| **Self-Reported Ongoing on Day of Survey** |  |  |
| Yes | 7289 | 45.0 |
| No | 8909 | 55.0 |

<sup>a</sup> The ARTnet survey allows participants to report details on up to 5 of the most recent sexual male partners within the past 12 months of taking the survey.

Mean degree estimates varied by partnership type for both measurement approaches. Using the current method, total mean degree was 1.19 across partnership types: 0.45 for main and 0.74 for casual partnerships (**Figure 2**). Mean degree estimates ranged from 1.01 at the 12-month offset to 1.23 at the 1-month offset across all partnerships. Mean degree ranged from 0.38 (12-month offset) to 0.45 (0-month offset) for main partnerships and from 0.64 (12-month offset) to 0.78 (1-month offset) for casual partnerships.

**Figure 2.**
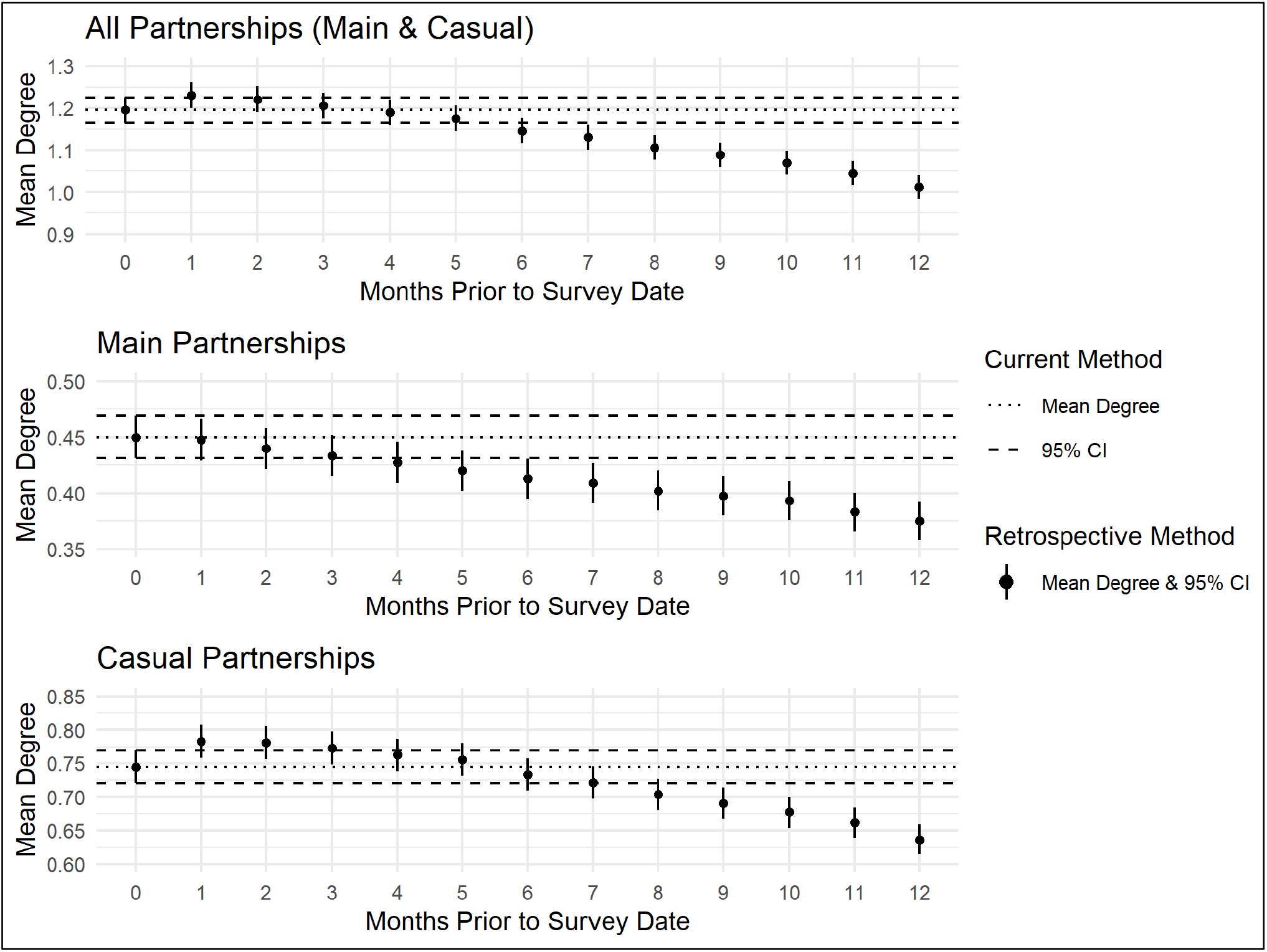
Comparison of mean degree calculated using the retrospective method from 0 to 12 months prior to survey date and mean degree calculated by the current method on the day of survey among all, main, and casual male sexual partnerships of 4,904 ARTnet participants.

The mean degrees estimated by the retrospective method decreased as the month offsets increased from 1 to 12 months (**Figure 2**). Mean degree estimated at the 0-month offset was the same as the mean degree estimated from the current method across all partnership types, which is expected given that the 0-month offset date is the same as the survey date for each participant. Mean degree of main partnerships from the current method (0.45) was most similar to estimates between 1- and 2-month offsets (0.45–0.44), while for casual partnerships, the current estimate (0.75) was most similar to mean degree estimated at 5- and 6-month offsets (0.76, 0.73). Overall mean degree estimated with the current method (1.19) was most similar to mean degree at the 4-month offset (1.19) across all partnerships.

When stratifying by having 5 or fewer or more than 5 partners in the past 12 months, the decreasing trend of mean degree was largely explained by participants who had more than 5 partners (**Figure 3**). Participants with more than 5 partners in the past year did not have the opportunity to share information on more than 5 of their partners, and therefore, the partnerships reported may be biased to the most recent partnerships. For main partnerships, there continued to be a decreasing trend of mean degree observed between the 1- and 12-month offsets (0.48 to 0.42) for participants with 5 or fewer partnerships in the past year; however, this did not impact the stability of the overall mean degree when restricted to participants with 5 or fewer partnerships. Mean degree remained stable from 0.83 at 1-month offset to 0.80 at 12-month offset for participants with 5 or fewer partners in the past year, but mean degree decreased from 1.83 at 1-month offset to 1.34 at 12-month offset for those with more than 5 partners. A similar pattern was found for casual mean degree. Among participants with 5 or fewer partners, casual mean degree was relatively stable from 0.35 at 1-month offset to 0.37 at 12-month offset. For participants with more than 5 partners, casual mean degree decreased from 1.44 at 1-month offset to 1.04 at 12-month offset. **Supplemental Table 1** describes mean degree estimates using both methods by participant and partnership characteristics for 0-, 3-, 6-, and 12-month offsets.

**Figure 3.**
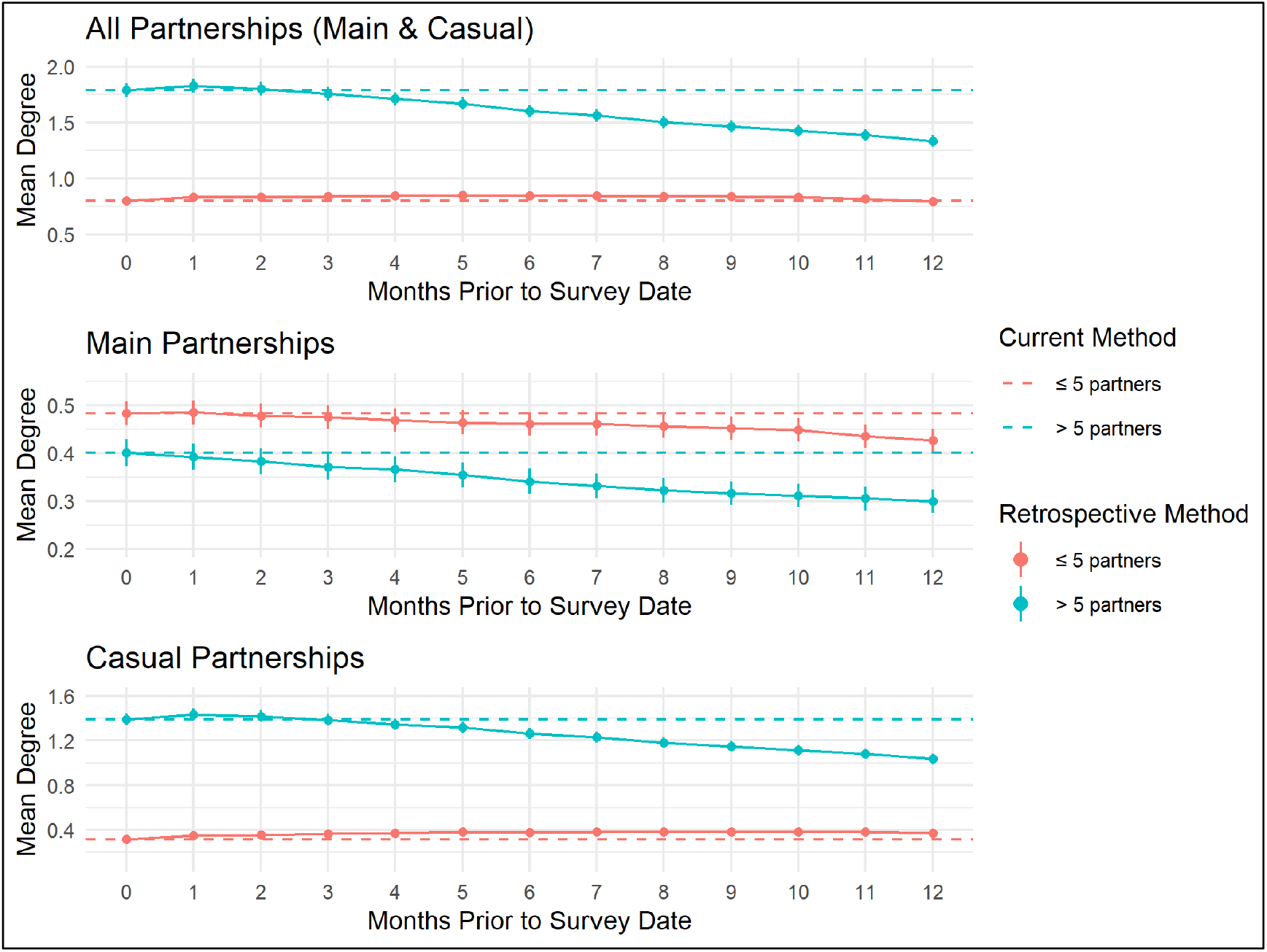
Comparison of mean degree calculated using the retrospective method from 0 to 12 months prior to survey date and mean degree calculated by the current method on the day of survey among all, main, and casual male sexual partnerships of 4,904 ARTnet participants (2017–2019) stratified by having 5 or fewer partners or more than 5 partners. ARTnet participants were only able to report on dates of first and last sex and whether partnerships were ongoing on a maximum of 5 partners, restricting calculation of mean degree to the 5 most recent partners.

Bivariable and multivariable linear regression further confirmed that the primary exposure of partnership truncation was the principal cause of the decreasing trend of retrospective mean degree estimates, particularly for casual partnerships. Race/ethnicity, age, and education were found to be potential confounders of the relationship between truncation of partnership data and the downward bias of mean degree in the retrospective method (**Table 2; Supplemental Table 2; Supplemental Figures 1–5**). The negative association between average change in degree and number of male partners in the past year remained after adjusting for demographics. The average change in main degree between 12- and 0-month offsets among participants with more than 5 partners in the past year compared to those with 5 or fewer partners was −0.04 (95% CI: −0.07, −0.02) in unadjusted analysis and −0.05 (95% CI: −0.08, −0.03) in adjusted analysis (**Table 2**). For casual partnerships among participants with more than 5 partners compared to 5 or fewer partners, the unadjusted average change in degree was −0.41 (95% CI: −0.46, −0.36) and adjusted average change in degree was −0.40 (95% CI: −0.45, −0.35).

**Table 2.** Bivariable linear regression results of average change in degree between 12 months and 0 months prior to survey date using the retrospective method for main and casual sexual partnerships of male participants (N = 4,904) in the ARTnet study (2017–2019)

|  | Main Partnerships |  |  | Casual Partnerships |  |  |
| --- | --- | --- | --- | --- | --- | --- |
|  | Estimate | 95% CI | F-Test P-value | Estimate | 95% CI | F-Test P-value |
| **Number of Male Partners<sup>a</sup>** |  |  | <0.01 |  |  | <0.01 |
| Intercept (≤ 5 Partners) | -0.06 | -0.07, -0.04 |  | 0.06 | 0.03, 0.09 |  |
| > 5 | -0.04 | -0.07, -0.02 |  | -0.41 | -0.46, -0.36 |  |
| **Race/Ethnicity** |  |  | 0.05 |  |  | 0.16 |
| Intercept (Black) | -0.02 | -0.07, 0.02 |  | -0.01 | -0.11, 0.10 |  |
| Hispanic | -0.08 | -0.13, -0.02 |  | -0.07 | -0.20, 0.04 |  |
| Other | -0.04 | -0.10, 0.01 |  | -0.09 | -0.22, 0.04 |  |
| White | -0.05 | -0.10, -0.01 |  | -0.11 | -0.23, 0.01 |  |
| **Age (continuous)** | 0.003 | 0.002, 0.004 | <0.01 | -0.003 | -0.005, -0.001 | <0.01 |
| **Age Category** |  |  | <0.01 |  |  | <0.01 |
| Intercept (15-24 years) | -0.15 | -0.17, -0.13 |  | -0.03 | -0.08, 0.01 |  |
| 25-34 years | 0.09 | 0.06, 0.12 |  | -0.07 | -0.14, -0.01 |  |
| 35-44 years | 0.09 | 0.05, 0.12 |  | -0.10 | -0.18, -0.02 |  |
| 45-54 years | 0.11 | 0.08, 0.15 |  | -0.15 | -0.23, -0.07 |  |
| 55-65 years | 0.13 | 0.09, 0.16 |  | -0.11 | -0.19, -0.03 |  |
| **Census Region** |  |  | 0.28 |  |  | 0.15 |
| Intercept (Northeast) | -0.06 | -0.08, -0.03 |  | -0.14 | -0.20, -0.08 |  |
| Midwest | -0.03 | -0.07, 0.00 |  | 0.07 | -0.01, 0.15 |  |
| South | -0.02 | -0.05, 0.01 |  | 0.05 | -0.02, 0.12 |  |
| West | -0.02 | -0.05, 0.02 |  | 0.00 | -0.08, 0.07 |  |
| **Highest Level of Education** |  |  | <0.01 |  |  | <0.01 |
| Intercept (High school or below) | -0.12 | -0.15, -0.09 |  | -0.06 | -0.13, 0.01 |  |
| Some college | 0.05 | 0.01, 0.08 |  | 0.03 | -0.05, 0.11 |  |
| College and above | 0.06 | 0.03, 0.09 |  | -0.11 | -0.18, -0.03 |  |
| **Annual Household Income** |  |  | 0.44 |  |  | <0.01 |
| Intercept (\$0 to \$19,999) | -0.08 | -0.11, -0.05 | | -0.01 | -0.08, 0.05 | |
| \$20,000 to \$39,999 | 0.00 | -0.04, 0.04 | | -0.08 | -0.17, 0.01 | |
| \$40,000 to \$74,999 | 0.00 | -0.04, 0.04 | | -0.07 | -0.15, 0.02 | |
| \$75,000 or more | 0.02 | -0.02, 0.05 | | -0.16 | -0.24, -0.08 | |

<sup>a</sup> The ARTnet survey allows participants to report details such as dates of first and last sex on up to 5 of the most recent sexual male partners within the past 12 months of taking the survey. Participants are also asked about the total number of male sexual partners they have had within the past 12 months but do not report details used to calculate mean degree on all of these partners.

In a sensitivity analysis on the influence of date imputation on retrospective mean degree, random date imputation had little effect on mean degree (**Supplemental Table 5**). When considering the minimum mean degree (sexual partnerships were not active during the entire month reported) and maximum mean degree (sexual partnerships were active during the entire month reported), there was an average percent change of 2–3%.

Twenty percent of one-time partnerships were considered active and ongoing on the day of the survey, but these partnerships were considered one-time partnerships and not included in mean degree estimation because participants had sex with these partners once. In a sensitivity analysis we redefined these partnerships as active, casual partnerships and considered a scenario where a proportion of “Don’t know” responses were “Yes.” These alternative definitions suggest our original definition may be underestimating the casual mean degree using the current method and the retrospective method at the 0-month offset (**Supplemental Table 6**). There was no effect on the retrospective mean degree beyond the 0-month offset.

## DISCUSSION

In this study, we compared the current and the retrospective methods of measuring mean degree in a sample of U.S. MSM. The two methods produced similar estimates of mean degree for participants with fewer past-year partners. With the retrospective method, mean degree estimates decreased as the retrospective measure of degree was calculated at more months prior to the survey date. This was due to truncated partnership data from the survey design: information used to calculate mean degree such as dates of first/last sex were collected on up to 5 of the most recent partners. Our results suggest that this downward bias leads to underestimation of degree, especially of casual partnerships, using the retrospective method when surveys have truncated partnership data. Underestimation of degree may impact projected transmission of HIV in simulated networks.^**23**,**24**^ The current method of measurement is less prone to bias from truncated partnership data but may have its own flaws.

With 2 in 5 ARTnet participants reporting more than 5 main, casual, and one-time male sexual partners in the past year, the truncation of partnership data had a major impact on mean degree estimation with the retrospective method. This was particularly true for casual partnerships, which were more prevalent compared to main partnerships in our sample. An exploration of a few cases with large differences in degree between 12- and 0-month offsets may offer some explanation for the downward bias (**Supplemental Figures 6–7)**. For example, in **Supplemental Figure 7**, study participant 4 reported 15 male sexual partners in the past 12 months but was only able to report on the 5 most recent partners, which were all considered ongoing on the day of survey but not active at 12 months prior to the survey date. This person’s degree would be 0 at the 12-month offset but 5 at the 0-month offset, resulting in a difference of −5. Since participants were asked to report on their most recent partnerships, participants with truncated partnership data were likely to only report on partnerships closer to the survey date before exhausting their responses, leading to underestimation of degree as month offsets increased. Furthermore, reporting on more one-time partnerships, which don’t contribute to mean degree, may censor main and casual partnerships in the past year and underestimate mean degree.

A direct comparison of the current method to the UNAIDS-recommended retrospective method at 6 months using ARTnet data shows that the percent of participants reporting more than 1 partner at both time points was similar and total, main, and casual mean degree was slightly smaller when estimated at the 6-month offset (**Supplemental Table 7**). The distribution of degree for both time points were also similar (**Supplemental Figure 8**). The retrospective method may still be a suitable alternative to the current method if there was no truncation of partnership data in the behavioral survey or if information on the active status of current partnerships was not collected and if mean degree is estimated within 6 months of the survey date.

The current or retrospective measurement methods have been applied to the estimation of partnership concurrency (degree of two or more), aligning generally with our current study. One study of young adults in the U.S. directly comparing reported partnership concurrency within the last 6 years through direct questioning and overlapping partnership dates found both methods reported a similar prevalence of concurrency.^**25**^ However, the agreement about concurrent partnerships between the two methods was only modest, with the authors suggesting that measuring concurrency using overlapping partnership dates was more likely to be complicated by missing or uninterpretable data. Another study of heterosexual partnerships in Malawi found that measuring concurrency using partnership dates underestimated self-reported concurrency due to difficulty of recalling partnership start and end dates and general underreporting of partnerships.^**26**^ The current method may be statistically preferable because both mean active degree and partnership age can be jointly estimated with typical distributional assumptions.^**20**,**27**^ On the other hand, study participants may not be able to accurately predict whether current partnerships will continue, leading to overestimation of mean active degree using the current method.^**11**^ The current method is not perfect, and bias adjustment or improvement of the measurement method is needed. Our analysis was unable to identify current partners that were truly active and ongoing because we did not collect follow-up data. Future studies may consider the use of longitudinal data where behavioral surveys administered at multiple time points collect data on a limited number of existing and new partners. This study design could evaluate the severity of the bias from truncation partnership data compared to bias from participants’ failure to predict ongoing partnerships.

### Limitations

There are some limitations to this study. First, only month and year of dates of first/last sex were reported for each partnership. Partnerships with start or end months that were the same as the month of the retrospective date from the date of survey could be considered ongoing or not ongoing depending on the exact day of first/last sex. To address this, we randomly imputed the day for the start and end dates to identify partnerships overlapping with the retrospective date. A sensitivity analysis found that the average percent change of mean degree for more extreme assumptions was minimal. Second, this study was limited to cross-sectional data, which may be useful because many studies estimate mean degree from cross-sectional data, but longitudinal studies may be able to describe unique biases from cross-sectional data such as the prediction of ongoing partnerships.^**11**^ Third, ARTnet coding decisions around one-time partnerships and “Do not know” responses for the question of whether a partnership is active and ongoing may lead to underestimation of mean degree using the current method or retrospective 0-month offset, which was confirmed through a sensitivity analysis. Given prior research that suggests MSM may be overconfident in predicting which sexual partnerships are ongoing,^**11**^ we used the original method as a conservative measure.

### Conclusion

Sexual mean active degree is an important measure in HIV and STI epidemiology. It should therefore be measured rigorously and estimated consistently across studies. Future network-based studies should justify their methods for mean active degree measurement with special consideration of how data is collected. Our analysis suggests that cross-sectional data with truncated partnership data may underestimate mean degree using the retrospective method, favoring the current method. However, improvements can further be made to the current method to address bias in predicting persistence of partnerships.^11^ The accuracy of the active degree estimates will have important impacts on the delivery of HIV/STI prevention interventions that target networks.

## Data Availability

All data produced in the present study are available upon reasonable request to the authors

https://github.com/EpiModel/Mean-Degree-Analysis

