## Supplemental Appendix for "Comparing Sexual Network Mean Active Degree Measurement Metrics among Men who have Sex with Men"

**Supplemental Table 1.** Comparison of mean degree estimated using the current method and retrospective method at 0, 3, 6, and 12 months prior to survey date stratified by partnership type and individual-characteristics among male participants in the ARTnet study (2017–2019)

| Variable | Level | ARTnet Participants (N) | Partnership Type | Current Method Mean Degree | Retrospective Mean Degree by Months Prior to Survey Date | | | |
|---|---|---|---|---|---|---|---|---|
| | | | | | 0 Months | 3 Months | 6 Months | 12 Months |
| **All** | | | Main | 0.450 | 0.450 | 0.433 | 0.413 | 0.375 |
| | | 4904 | Casual | 0.745 | 0.745 | 0.772 | 0.733 | 0.637 |
| **Total Partners** | | | Main | 0.483 | 0.483 | 0.475 | 0.461 | 0.426 |
| | 5 or fewer | 2939 | Casual | 0.315 | 0.315 | 0.365 | 0.380 | 0.371 |
| | | | Main | 0.401 | 0.401 | 0.372 | 0.341 | 0.300 |
| | 6 or more | 1962 | Casual | 1.390 | 1.390 | 1.384 | 1.263 | 1.036 |
| **Race/ Ethnicity** | | | Main | 0.316 | 0.316 | 0.323 | 0.323 | 0.293 |
| | Black | 266 | Casual | 0.808 | 0.808 | 0.883 | 0.887 | 0.801 |
| | | | Main | 0.451 | 0.451 | 0.429 | 0.399 | 0.354 |
| | Hispanic | 676 | Casual | 0.691 | 0.691 | 0.743 | 0.704 | 0.609 |
| | | | Main | 0.410 | 0.410 | 0.401 | 0.394 | 0.344 |
| | Other | 439 | Casual | 0.699 | 0.699 | 0.768 | 0.697 | 0.604 |
| | | | Main | 0.465 | 0.465 | 0.446 | 0.424 | 0.389 |
| | White | 3523 | Casual | 0.756 | 0.756 | 0.770 | 0.732 | 0.634 |
| **Census Region** | | | Main | 0.461 | 0.461 | 0.435 | 0.409 | 0.372 |
| | Midwest | 994 | Casual | 0.722 | 0.722 | 0.759 | 0.718 | 0.653 |
| | | | Main | 0.444 | 0.444 | 0.432 | 0.412 | 0.389 |
| | Northeast | 882 | Casual | 0.805 | 0.805 | 0.840 | 0.781 | 0.667 |
| | | | Main | 0.457 | 0.457 | 0.439 | 0.421 | 0.379 |
| | South | 1782 | Casual | 0.732 | 0.732 | 0.753 | 0.728 | 0.640 |
| | | | Main | 0.435 | 0.435 | 0.425 | 0.404 | 0.363 |
| | West | 1246 | Casual | 0.738 | 0.738 | 0.764 | 0.719 | 0.598 |
| **Age** | | | Main | 0.415 | 0.415 | 0.384 | 0.339 | 0.265 |
| | 15-24 | 1324 | Casual | 0.401 | 0.401 | 0.433 | 0.425 | 0.369 |
| | | | Main | 0.517 | 0.517 | 0.500 | 0.483 | 0.455 |
| | 25-34 | 1268 | Casual | 0.573 | 0.573 | 0.632 | 0.577 | 0.468 |
| | | | Main | 0.507 | 0.507 | 0.491 | 0.474 | 0.447 |
| | 35-44 | 694 | Casual | 0.889 | 0.889 | 0.915 | 0.866 | 0.758 |
| | | | Main | 0.449 | 0.449 | 0.445 | 0.437 | 0.413 |
| | 45-54 | 833 | Casual | 1.091 | 1.091 | 1.079 | 1.030 | 0.909 |
| | | | Main | 0.352 | 0.352 | 0.345 | 0.343 | 0.329 |
| | 55-65 | 785 | Casual | 1.106 | 1.106 | 1.120 | 1.073 | 0.964 |
| **Education** | | | Main | 0.372 | 0.372 | 0.354 | 0.310 | 0.248 |
| | High school or below | 616 | Casual | 0.526 | 0.526 | 0.542 | 0.544 | 0.469 |
| | | | Main | 0.433 | 0.433 | 0.410 | 0.388 | 0.357 |
| | Some college | 1519 | Casual | 0.668 | 0.668 | 0.736 | 0.725 | 0.641 |
| | | | Main | 0.478 | 0.478 | 0.465 | 0.450 | 0.414 |
| | College and above | 2742 | Casual | 0.837 | 0.837 | 0.845 | 0.783 | 0.674 |
| **Income** | | | Main | 0.345 | 0.345 | 0.342 | 0.331 | 0.268 |
| | $0 to $19,999 | 605 | Casual | 0.545 | 0.545 | 0.643 | 0.617 | 0.531 |
| | | | Main | 0.442 | 0.442 | 0.428 | 0.392 | 0.366 |
| | $20,000 to $39,999 | 867 | Casual | 0.694 | 0.694 | 0.724 | 0.686 | 0.599 |
| | | | Main | 0.429 | 0.429 | 0.409 | 0.385 | 0.351 |
| | $40,000 to $74,999 | 1258 | Casual | 0.741 | 0.741 | 0.788 | 0.744 | 0.660 |
| | | | Main | 0.517 | 0.517 | 0.505 | 0.492 | 0.458 |
| | $75,000 or more | 1686 | Casual | 0.870 | 0.870 | 0.855 | 0.809 | 0.698 |

**Supplemental Table 2.** Characteristics of ARTnet study participants (N = 4,904; 2017–2019) by total number of sexual partners in the past 12 months. Total number of sexual partners set at threshold value of 5 for maximum number of sexual partners ARTnet participants can report dates of first and last sex for mean degree estimation.

| | ARTnet Participants, N (Column %) | |
|---|---|---|
| **Characteristic** | ≤5 Partners | >5 Partners |
| **Race/Ethnicity** | | |
| Black (Non-Hispanic) | 174 (5.9) | 92 (4.7) |
| Hispanic | 406 (13.8) | 268 (13.7) |
| Other (Non-Hispanic) | 256 (8.7) | 183 (9.3) |
| White (Non-Hispanic) | 2103 (71.6) | 1419 (72.3) |
| **Age Category** | | |
| 15–24 | 893 (30.4) | 431 (22.0) |
| 25–34 | 774 (26.3) | 494 (25.2) |
| 35–44 | 382 (13.0) | 312 (15.9) |
| 45–54 | 452 (15.4) | 381 (19.4) |
| 55–65 | 438 (14.9) | 344 (17.5) |
| **Education** | | |
| High school or below | 447 (15.3) | 169 (8.7) |
| Some college | 970 (33.2) | 548 (28.1) |
| College and above | 1506 (51.5) | 1234 (63.2) |

**Supplemental Table 3.** Cross-tabulation of the number of ARTnet study participants (N = 4,904; 2017–2019) with a total of 5 or fewer male sexual partners more than 5 partners with the total number of sexual partnerships for which participants provided details such as dates of first and last sex (maximum of 5 due to survey design)

| **Total Number of Male Sexual Partners in the Past 12 Months** | **Total Number of Male Sexual Partnerships Reported on in ARTnet (Maximum of 5)** | | | | | |
|---|---|---|---|---|---|---|
| | **0** | **1** | **2** | **3** | **4** | **5** |
| **≤ 5 Partners** | 234 | 1062 | 557 | 444 | 319 | 310 |
| **> 5 Partners** | 0 | 0 | 2 | 2 | 1 | 1970 |

**Supplemental Table 4.** Bivariable Linear Regression Results of Average Change in Degree of Main and Casual Partnerships Between 12-Month and 0-Month Offsets by total number of partners in the past 12 months

| | Main Partnerships | | Casual Partnerships | |
|---|---|---|---|---|
| **Total Partners** | **Estimate** | **95% CI** | **Estimate** | **95% CI** |
| **Intercept (0)** | 0.00 | -0.05, 0.05 | 0.00 | -0.11, 0.11 |
| **1** | -0.07 | -0.13, -0.02 | 0.03 | -0.09, 0.15 |
| **2** | -0.05 | -0.10, 0.01 | 0.10 | -0.03, 0.23 |
| **3** | -0.02 | -0.08, 0.04 | 0.07 | -0.06, 0.21 |
| **4** | -0.13 | -0.19, -0.06 | 0.08 | -0.06, 0.22 |
| **5** | -0.04 | -0.10, 0.03 | 0.02 | -0.12, 0.17 |
| **> 5** | -0.10 | -0.15, -0.05 | -0.36 | -0.47, -0.24 |

**Supplemental Table 5.** Sensitivity analysis of the influence of date imputation on mean degree estimated using the retrospective method from 0 to 12 months prior to survey date by partnership type (total, main, and casual) for male-male sexual partnerships in the ARTnet study (N = 4,904; 2017–2019). “Min” refers to the minimum mean degree based on the assumption that a sexual partnership is not active during the entire month reported for the dates of first and last sex. “Obs” refers to the observed mean degree in the analysis based on random date imputation where some sexual partnerships may be active and some sexual partnerships may not. “Max” refers to the maximum mean degree based on the assumption that a sexual partnership is active for the entire month reported for the dates of first and last sex.

| **Month** | **Total (Min)** | **Total (Obs)** | **Total (Max)** | **Max-Min** | **Main (Min)** | **Main (Obs)** | **Main (Max)** | **Max-Min** | **Casual (Min)** | **Casual (Obs)** | **Casual (Max)** | **Max-Min** |
|---|---|---|---|---|---|---|---|---|---|---|---|---|
| 0 | | 1.19 | 1.20 | | | 0.45 | 0.45 | | | 0.74 | 0.75 | |
| 1 | 1.18 | 1.23 | 1.27 | 0.09 | 0.44 | 0.45 | 0.45 | 0.02 | 0.74 | 0.78 | 0.82 | 0.07 |
| 2 | 1.17 | 1.22 | 1.26 | 0.09 | 0.43 | 0.44 | 0.44 | 0.01 | 0.74 | 0.78 | 0.82 | 0.08 |
| 3 | 1.16 | 1.21 | 1.24 | 0.08 | 0.42 | 0.43 | 0.44 | 0.02 | 0.74 | 0.77 | 0.80 | 0.06 |
| 4 | 1.15 | 1.19 | 1.22 | 0.07 | 0.42 | 0.43 | 0.43 | 0.01 | 0.73 | 0.76 | 0.79 | 0.05 |
| 5 | 1.14 | 1.18 | 1.20 | 0.07 | 0.41 | 0.42 | 0.43 | 0.01 | 0.73 | 0.76 | 0.78 | 0.05 |
| 6 | 1.12 | 1.15 | 1.17 | 0.06 | 0.41 | 0.41 | 0.42 | 0.01 | 0.71 | 0.73 | 0.76 | 0.05 |
| 7 | 1.09 | 1.13 | 1.15 | 0.06 | 0.40 | 0.41 | 0.41 | 0.01 | 0.69 | 0.72 | 0.74 | 0.05 |
| 8 | 1.07 | 1.11 | 1.13 | 0.05 | 0.39 | 0.40 | 0.41 | 0.01 | 0.68 | 0.70 | 0.72 | 0.04 |
| 9 | 1.06 | 1.09 | 1.10 | 0.04 | 0.39 | 0.40 | 0.40 | 0.01 | 0.67 | 0.69 | 0.70 | 0.03 |
| 10 | 1.04 | 1.07 | 1.09 | 0.04 | 0.38 | 0.39 | 0.40 | 0.01 | 0.66 | 0.68 | 0.69 | 0.03 |
| 11 | 1.02 | 1.04 | 1.07 | 0.05 | 0.38 | 0.38 | 0.39 | 0.01 | 0.64 | 0.66 | 0.68 | 0.03 |
| 12 | 0.99 | 1.01 | 1.03 | 0.04 | 0.37 | 0.38 | 0.38 | 0.01 | 0.62 | 0.64 | 0.65 | 0.03 |
| Average | | | | 0.0610 | | | | 0.0132 | | | | 0.0477 |
| Average percent change +/- | | | | 3% | | | | 2% | | | | 3% |

**Supplemental Table 6.** Sensitivity analysis of casual mean degree for alternative definitions of a sexual partnership's ongoing status for ARTnet participants (N = 4,904; 2017–2019): 1) alternative assumption that partnerships marked ongoing on day of survey are all considered ongoing even if partners have only had sex with the partner once, which changes some one-time partnerships to casual partnerships; and 2) imputation of "Don't know" responses to ongoing status variable on day of survey. The original assumption used in the analysis is if participants had sex with a partner only once, then that sexual partnership is considered a one-time partnership even if the participant believes they will have sex with that person again. These assumptions only affect casual mean degree.

| Active Indicator Status & Partnership Type Definition | Current Method Casual Mean Degree | Retrospective Casual Mean Degree (Month 0) | Retrospective Casual Mean Degree (Month 1) |
|---|---|---|---|
| **Casual Partners** | | | |
| Original[a] | 0.74 | 0.75 | 0.79 |
| Alternative[b] | 1.03 | 1.04 | 0.79 |
| Imputed[c] | 1.00 | 1.04 | 0.79 |
| **Total Partners** | | | |
| Original | 1.19 | 1.2 | 1.23 |
| Alternative | 1.49 | 1.45 | 1.23 |
| Imputed | 1.48 | 1.49 | 1.23 |

[a] Original: if participant only had sex once with partner, then the sexual partnership is considered a one-time partnership even if the participant expects to have sex with them again

[b] Alternative: all partnerships where the participant expects to have sex with the partner again are considered ongoing even if partners only had sex with each other once by the survey date

[c] Imputed: "Don't know" response to whether a partnership was active and ongoing was randomly imputed to ongoing by partnership type: 1) for casual partnerships, a probability of 0.5 was used to change "Don't know" responses to ongoing; 2) for one-time partnerships, a probability of 0.1 was used to change "Don't know" responses to ongoing, and using the alternative definition, these partnerships were then considered active and ongoing

**Supplemental Table 7.** Comparison of the cross-sectional percentage of ARTnet participants (N = 4,904; 2017–2019) reporting more than one partner with the current method and UNAIDS-recommended retrospective method at 6 months prior to the survey date

| Metric | Percent of participants reporting more than 1 partner (95% CI) | Overall Mean Degree (95% CI) | Main Mean Degree | Casual Mean Degree |
|---|---|---|---|---|
| Current method | 26.4 (25.2, 27.6) | 1.19 (1.16, 1.23) | 0.45 (0.43, 0.47) | 0.75 (0.72, 0.77) |
| UNAIDS 6-month retrospective | 26.3 (25.1, 27.5) | 1.15 (1.12, 1.18) | 0.42 (0.39, 0.43) | 0.73 (0.71, 0.74) |

**Supplemental Figure 1.** Comparison of mean degree calculated using the retrospective method from 0 to 12 months prior to survey date and mean degree calculated by the current method on the day of survey among all, main, and casual male sexual partnerships of 4,904 ARTnet participants (2017–2019) stratified by race/ethnicity.

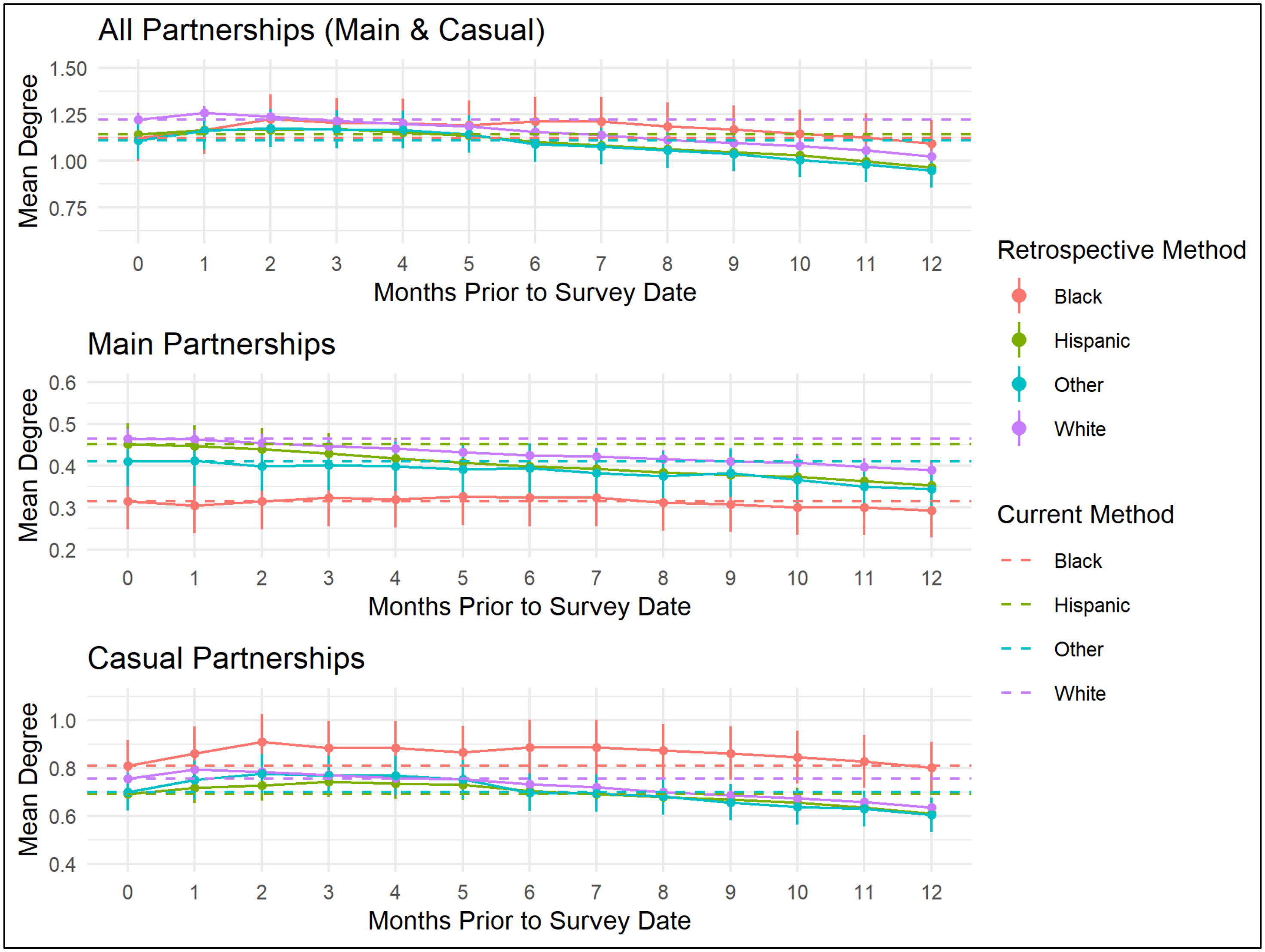

**Supplemental Figure 2.** Comparison of mean degree calculated using the retrospective method from 0 to 12 months prior to survey date and mean degree calculated by the current method on the day of survey among all, main, and casual male sexual partnerships of 4,904 ARTnet participants (2017–2019) stratified by age group.

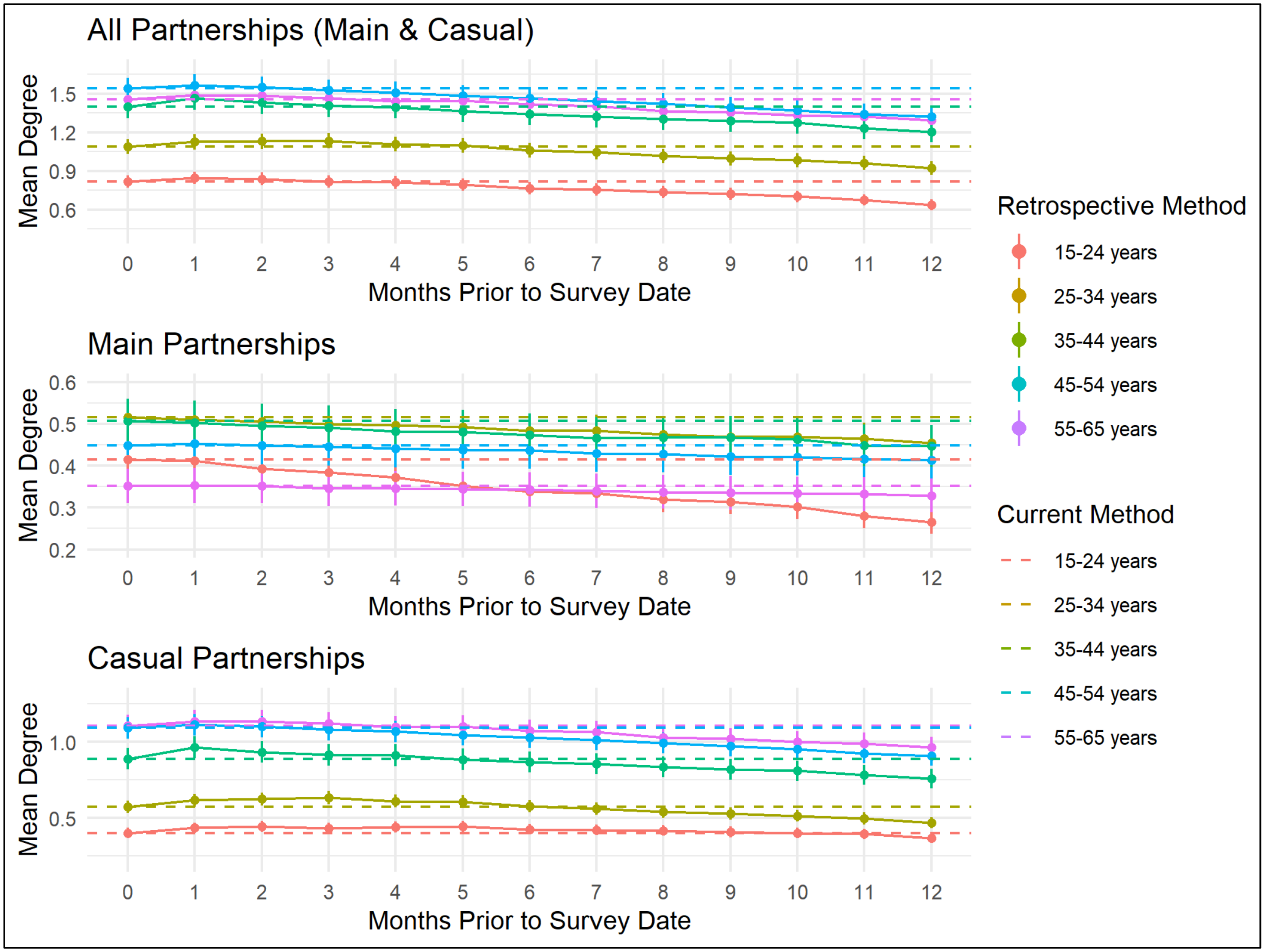

**Supplemental Figure 3.** Comparison of mean degree calculated using the retrospective method from 0 to 12 months prior to survey date and mean degree calculated by the current method on the day of survey among all, main, and casual male sexual partnerships of 4,904 ARTnet participants (2017–2019) stratified by census region.

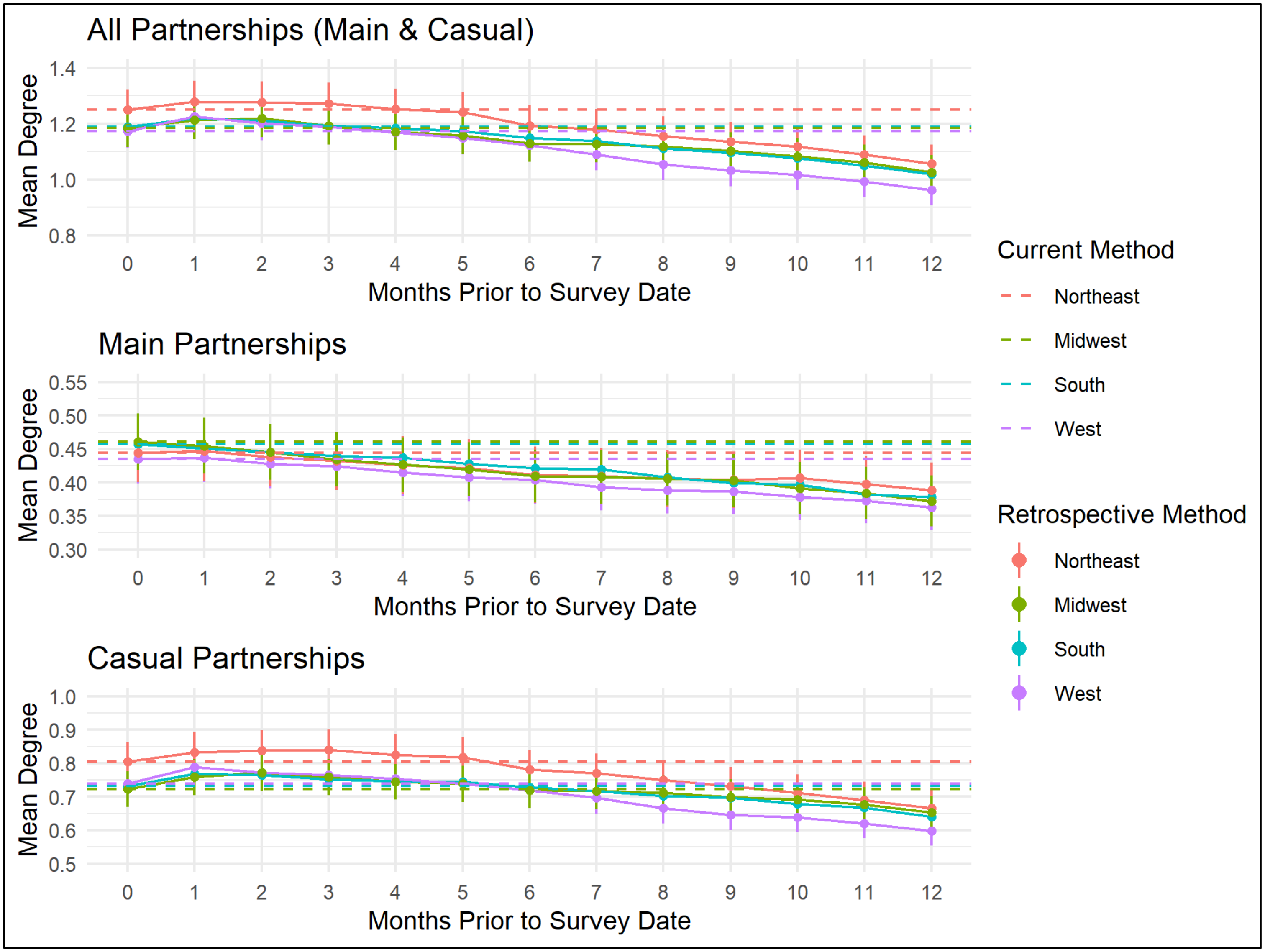

**Supplemental Figure 4.** Comparison of mean degree calculated using the retrospective method from 0 to 12 months prior to survey date and mean degree calculated by the current method on the day of survey among all, main, and casual male sexual partnerships of 4,904 ARTnet participants (2017–2019) stratified by highest level of education.

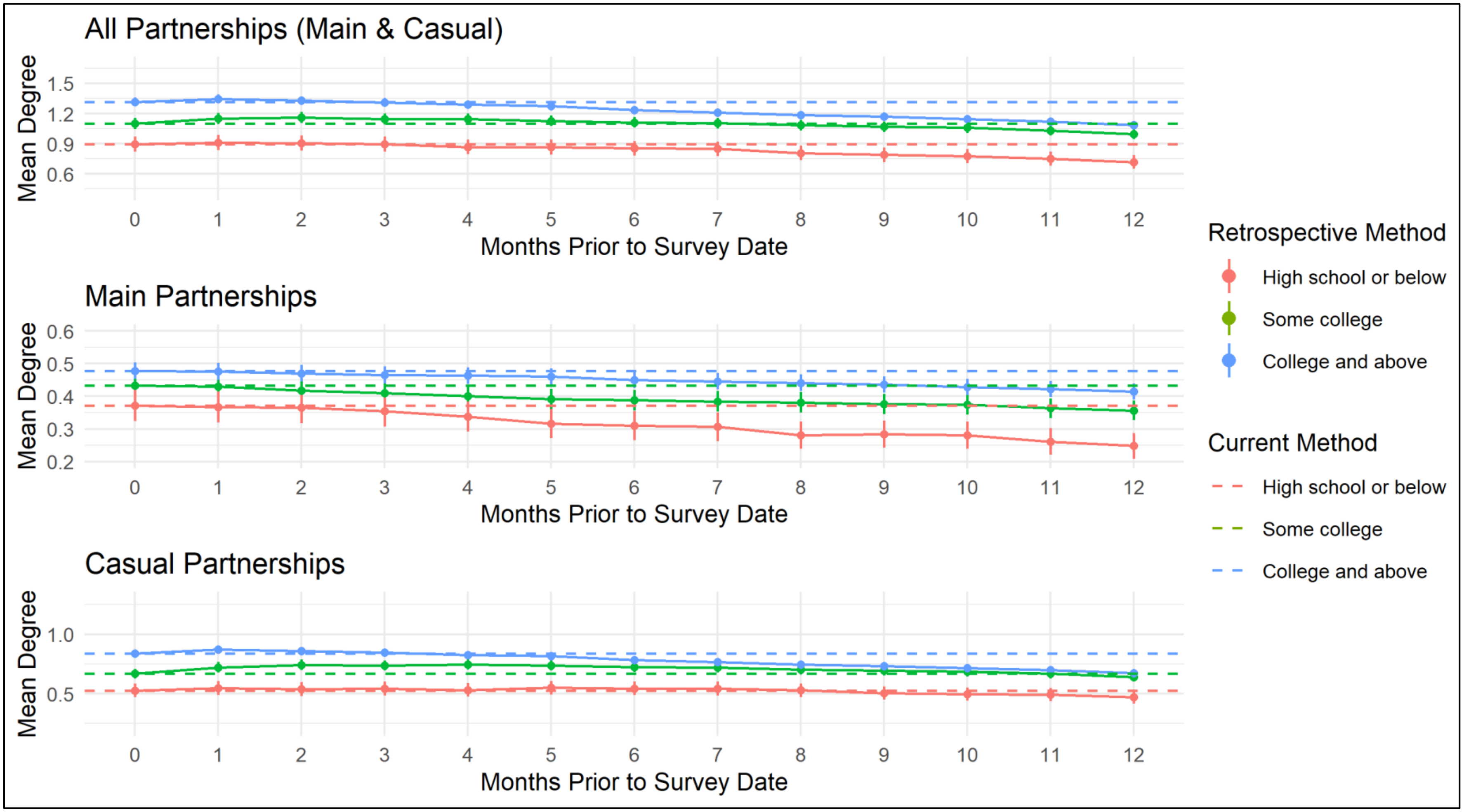

**Supplemental Figure 5.** Comparison of mean degree calculated using the retrospective method from 0 to 12 months prior to survey date and mean degree calculated by the current method on the day of survey among all, main, and casual male sexual partnerships of 4,904 ARTnet participants (2017–2019) stratified by income.

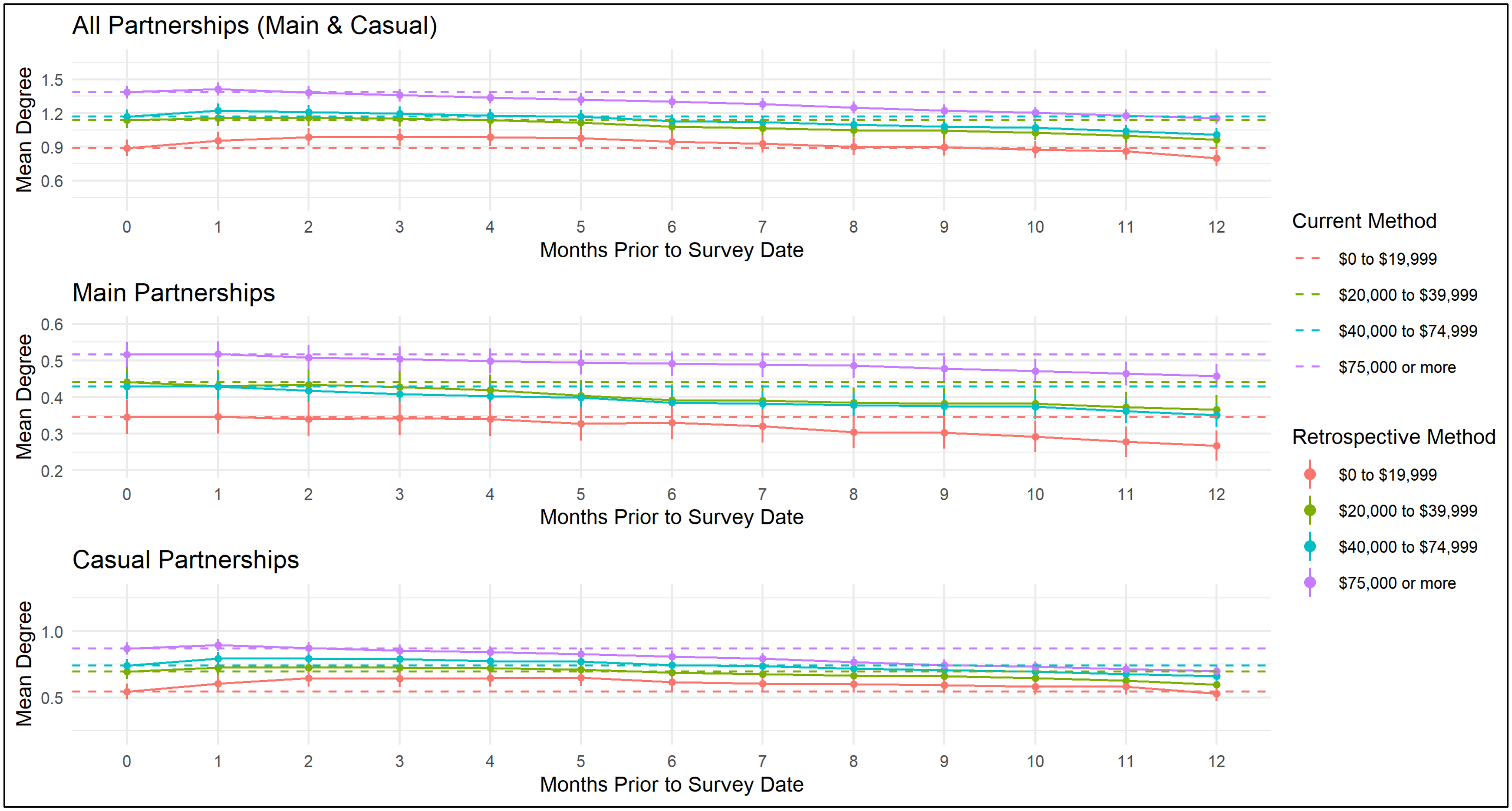

**Supplemental Figure 6.** Sexual partnership details (ongoing, partner type, partnership duration within the past 12 months) for selected ARTnet participants (N = 4,904; 2017–2019) with large differences in degree between 12- and 0- months prior to survey date for main partnerships.

| | | | | | | Months Prior to Survey Date | | | | | | | | | | | | |
|---|---|---|---|---|---|---|---|---|---|---|---|---|---|---|---|---|---|---|
| **Study Participant** | **Difference in Main Partnership Degree Between 12- and 0- months Prior to Survey Date** | **Total Number of Partners in the Past Year** | **Partner** | **Ongoing** | **Partner Type** | **0** | **1** | **2** | **3** | **4** | **5** | **6** | **7** | **8** | **9** | **10** | **11** | **12** |
| **1** | **-2** | **20** | 1 | No | One-Time | | | | | | | | | | | | | |
| | | | 2 | Yes | One-Time | | | | | | | | | | | | | |
| | | | 3 | Yes | Main | | | | | | | | | | | | | |
| | | | 4 | Yes | Main | | | | | | | | | | | | | |
| | | | 5 | No | One-Time | | | | | | | | | | | | | |
| **2** | **-2** | **31** | 1 | Yes | One-Time | | | | | | | | | | | | | |
| | | | 2 | Yes | Main | | | | | | | | | | | | | |
| | | | 3 | No | One-Time | | | | | | | | | | | | | |
| | | | 4 | No | One-Time | | | | | | | | | | | | | |
| | | | 5 | Yes | Main | | | | | | | | | | | | | |
| **3** | **2** | **3** | 1 | No | Main | | | | | | | | | | | | | |
| | | | 2 | No | Main | | | | | | | | | | | | | |
| | | | 3 | Yes | One-Time | | | | | | | | | | | | | |
| **4** | **3** | **6** | 1 | No | Main | | | | | | | | | | | | | |
| | | | 2 | No | Main | | | | | | | | | | | | | |
| | | | 3 | No | One-Time | | | | | | | | | | | | | |
| | | | 4 | No | One-Time | | | | | | | | | | | | | |
| | | | 5 | No | Main | | | | | | | | | | | | | |
| **5** | **-2** | **40** | 1 | Yes | Main | | | | | | | | | | | | | |
| | | | 2 | No | Casual | | | | | | | | | | | | | |
| | | | 3 | No | One-Time | | | | | | | | | | | | | |
| | | | 4 | No | One-Time | | | | | | | | | | | | | |
| | | | 5 | Yes | Main | | | | | | | | | | | | | |

**Supplemental Figure 7.** Sexual partnership details (ongoing, partner type, partnership duration within the past 12 months) for selected ARTnet participants (N = 4,904; 2017–2019) with large differences in degree between 12- and 0- months prior to survey date for casual partnerships.

| Study Participant | Difference in Casual Partnership Degree Between 12- and 0- months Prior to Survey Date | Total Number of Partners in the Past Year | Partner | Ongoing | Partner Type | Months Prior to Survey Date | | | | | | | | | | | | |
|---|---|---|---|---|---|---|---|---|---|---|---|---|---|---|---|---|---|---|
| | | | | | | 0 | 1 | 2 | 3 | 4 | 5 | 6 | 7 | 8 | 9 | 10 | 11 | 12 |
| **1** | **-4** | **5** | 1 | Yes | Casual | | | | | | | | | | | | | |
| | | | 2 | Yes | Casual | | | | | | | | | | | | | |
| | | | 3 | Yes | Casual | | | | | | | | | | | | | |
| | | | 4 | Yes | Casual | | | | | | | | | | | | | |
| | | | 5 | No | Casual | | | | | | | | | | | | | |
| **2** | **5** | **6** | 1 | No | Casual | | | | | | | | | | | | | |
| | | | 2 | No | Casual | | | | | | | | | | | | | |
| | | | 3 | No | Casual | | | | | | | | | | | | | |
| | | | 4 | No | Casual | | | | | | | | | | | | | |
| | | | 5 | No | Casual | | | | | | | | | | | | | |
| **3** | **4** | **5** | 1 | No | One-Time | | | | | | | | | | | | | |
| | | | 2 | No | Casual | | | | | | | | | | | | | |
| | | | 3 | No | Casual | | | | | | | | | | | | | |
| | | | 4 | No | Casual | | | | | | | | | | | | | |
| | | | 5 | No | Casual | | | | | | | | | | | | | |
| **4** | **-5** | **15** | 1 | Yes | Casual | | | | | | | | | | | | | |
| | | | 2 | Yes | Casual | | | | | | | | | | | | | |
| | | | 3 | Yes | Casual | | | | | | | | | | | | | |
| | | | 4 | Yes | Casual | | | | | | | | | | | | | |
| | | | 5 | Yes | Casual | | | | | | | | | | | | | |
| **5** | **-4** | **300** | 1 | Yes | Casual | | | | | | | | | | | | | |
| | | | 2 | Yes | Casual | | | | | | | | | | | | | |
| | | | 3 | Yes | Casual | | | | | | | | | | | | | |
| | | | 4 | Yes | Casual | | | | | | | | | | | | | |
| | | | 5 | Yes | Casual | | | | | | | | | | | | | |

**Supplemental Figure 8.** Distribution of degree with the current method and the retrospective method at 6 months prior to the survey date among ARTnet study participants (N = 4,904; 2017–2019)

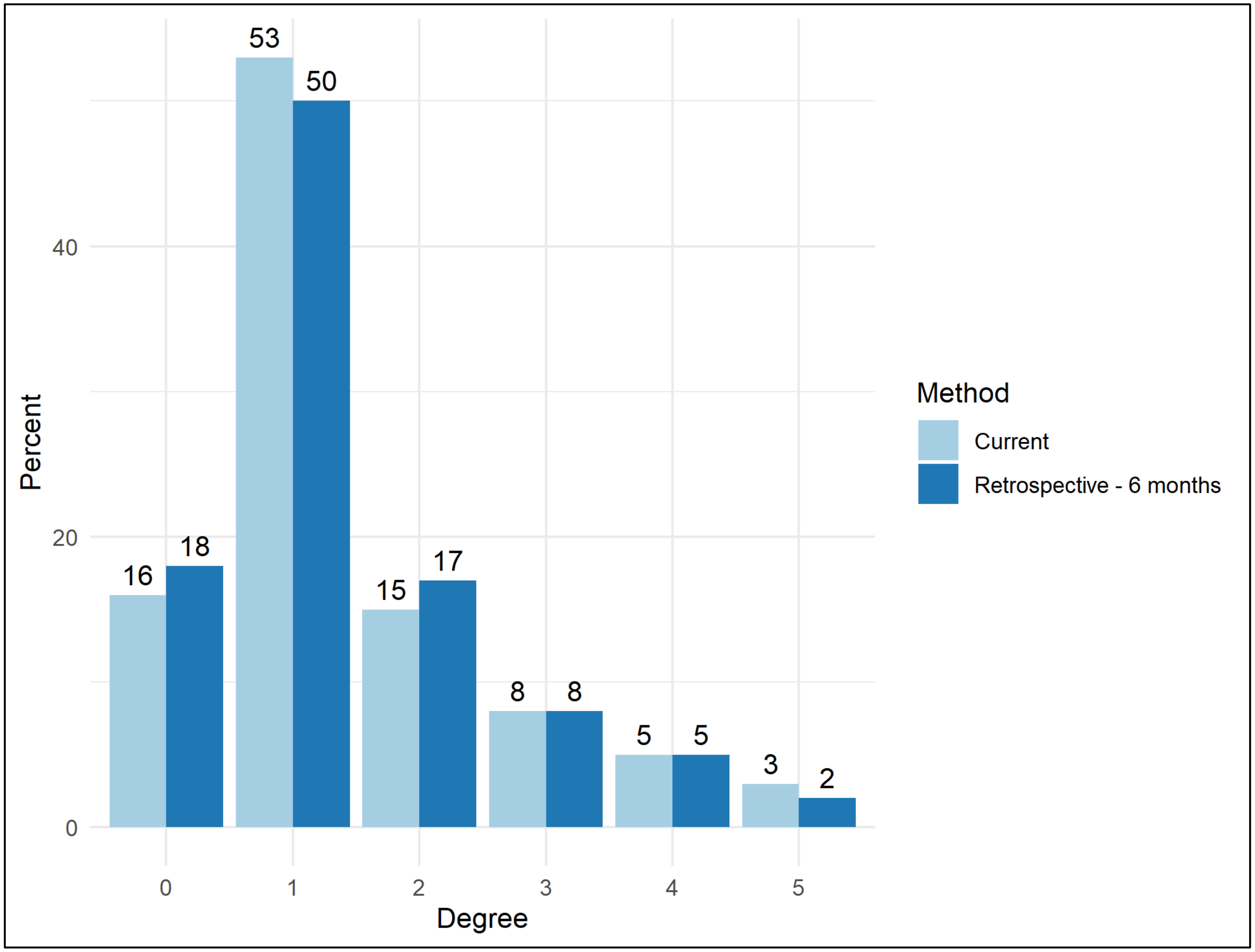
